# Retrospective Clinical Surveillance Measuring Healthcare Associated Infection (HAI) Rates Pre-and Post-Inclusion of Novel Silver Ion Antimicrobial Textile Intervention in an Infection Control Program

**DOI:** 10.1101/2020.12.09.20246702

**Authors:** Priya Balachandran, Kavita Mathur, J. Trees Ritter

## Abstract

Healthcare linens act as a vector of microbial transmission through use, storage and handling. In this retrospective multi-year, multi-site study, the impact of an infection prevention program, that included an automated silver ion-based antimicrobial laundry treatment, was studied. A composite reduction of 42% in healthcare associated infection (HAI) rates was observed, with the biggest reductions associated with CAUTI and CDI rates. Although further study is needed to better understand the exact contribution of such an intervention towards prevention of HAIs, ionic silver treatment of healthcare textiles may prove to be a useful tool in HAI reduction strategies.

## INTRODUCTION

Healthcare textiles are recognized as an epidemiologically important environmental surface implicated in outbreaks caused by pathogens within the hospital environment^1-4^. A recent environmental sampling study for 13 confirmed COVID-19 patients found extensive contamination of patients’ surroundings including pillows and bedsheets^5^. Microbial contamination of healthcare textiles occurs at mutliple points, each contributing in varying degrees to the spread of infections. Linens become contaminated while in use by patients and healthcare workers, due to intimate contact with the patients’ skin, which constantly sheds pathogen laden skin scales and hair fibers^1,3,4^. Similarly, ambulatory patients track pathogens on the soles of their feet/socks back into the bed ^6^. In hospitals where bed linens are not changed daily, contamination poses an infection risk to the patient who has indwelling devices such as urinary and vascular catheters which provide a direct portal of entry for pathogens^7^. Privacy curtains are another type of soft surface/textile in patient rooms that commonly become contaminated due to frequent handling. Contamination, especially on the grab area of the curtains, acts as a source of pathogen transmission from patient to patient via healthcare worker hands^1^. Another source of contamination occurs when clean linens are handled during transportion and storage^2^. Finally, pathogens such as *Clostridoides difficile* remain after laundering if stringent disinfection parameters are not met, subjecting the next patient to infection risk^8^.

Studies have demonstrated that incorporating metal ion-based antimicrobials in soft and hard surfaces in hospitals reduce healthcare-associated pathogens and associated infections^3,4^. For example, the use of copper-threaded/embedded fabrics in healthcare settings has achieved reductions in the pathogens that cause HAIs, such as *Staphylococcus aureus* (*S. aureus*), *Escherichia coli*, norovirus and other mutli-drug resistant organisms (MDROs)^3^. While effective, these threaded/embedded textiles require purchase of new, specialized inventory which can be costly to implement, maintain and replace when inventory is lost or degrades over time.

Our previous work found that hospital textiles treated with ionic silver during each laundry cycle resulted in over an 80% reduction in post-laundry levels of aerobic bacterial counts, *Staphylococcus aureus* and methicillin resistant *Staphylococcus aureus* (MRSA), pre- and post-patient use ^4^. Ionic silver has broad spectrum antibacterial, antifungal and antiviral activity ^8^. In this follow-on study continued at the same community hospital locations, the impact of the same silver-ion treatment of textiles was evaluated on HAI rates when implemented as a component of an infection prevention program to provide textile-mediated immediate and residual antimicrobial activity.

## METHODS

A retrospective, non-randomized before-and-after control study was designed to assess the efficacy of a silver ion-based laundry additive on HAI rates over an 18-month period (January 2016 to June 2017) and a 30-months period (January 2016 to June 2018). The study was conducted at five community hospitals located within the same hospital system, three of which served as treatment hospitals. All three treatment hospitals have similar patient acuity levels, demographics and are serviced by the same laundry facility accredited by the Healthcare Laundry Accreditation Council (HLAC). Hospital 1 had 286 available beds with a 67% occupancy rate, Hospital 2 had 112 available beds with a 67.5 % occupancy rate and Hospital 3 had 35 available beds with a 67% occupancy rate. All three hospitals had average length of stay of 4 days.

Two additional hospitals within the same hospital system, with similar patient acuity levels, demographics and infection prevention programs, served as control sites during 18 of the 30-month baseline period (“before”), and 18 of the 30-month after intervention period (“after”). Control hospital 1 had 155 available beds with a 46% occupancy rate, Control hospital 2 had 240 available beds with a 52% occupancy rate. Both hospitals had an average length of stay 4 days. At 18 months into the study period, based on the success of the study hospitals, the control hospitals opted to include silver ion laundry treatment for their infection control program and discontinued serving as control sites.

In January 2016, the silver ion dosing unit installed at the contract laundry facility servicing the three treatment hospitals was turned on and the treatment of laundry was monitored via cloud-based remote telemetry system (SilvaClean by Applied Silver Reg. No. 90335-1). During the intervention period the hospitals received antimicrobial treated flat patient bed sheets, fitted sheets, pillowcases and patient gowns. During the same period the control hopsitals received sheets, pillowcases and patient gowns laundered in the usual process without the addition of any silver ion antimicrobial treatment.

The silver ion treatment requires no change in laundry facility staffing or workflow. The ionic silver solution is introduced during the laundry rinse cycle in an automated fashion.

There were no reported changes in any other infection prevention (IP) products or practices in any of the hospitals during the study. The IP products and practices in place at each of the study and control hospitals were aligned with professional clinical guidelines from the Center for Disease Control and Prevention (CDC) and the Association for Infection Control and Epidemiology (APIC). HAI surveillance was conducted retrospectively for catheter associated urinary tract infection (CAUTI), central line associated bloodstream infection (CLABSI), *Clostridium difficile* infection (CDI) and surgical site infection (SSI). HAI definitions were based on the CDC’s National Health and Safety Network (NHSN) program.

Baseline HAI rates were documented for 30 months from July 2013 to December 2015. HAI surveillance continued during the 30-month intervention period from January 2016 to June 2018, while the silver ion-based laundry additive was in use. Infection rates were normalized with a denominator of 10,000 patient days. Statistical analysis included use of the Fisher exact test.

## RESULTS AND DISCUSSIONS

In a prior peer-reviewed published study, treatment of incumbent linens with silver ions during laundering resulted in statistically significant reductions of > 80% in the levels of total bacteria, *S. aureus* and MRSA, both pre and post patient use, as a result of residual antimicrobial effect^4^. This current follow-on study shows that an infection prevention bundle strategy at the three community hospitals within a hospital system, which included this seamless, automated silver ion laundry technology, reduced overall healthcare associated infection rates by 42% after 30 months of implementation.

In the current study, the impact of the silver-ion based laundry treatment on individual HAI types varied. Although all HAI types were not measured, of those tracked during the study (CDI, CAUTI, CLABSI and SSI) clinically significant reductions were seen in CDI and CAUTI. There was a statistically significant (50%) reduction in CDI from 4.7 to 2.4/10,000 patient days (p=0.001), and a 41% reduction (not statistically significant) in CAUTI from 8.8 to 5.2/10,000 patient days (p=0.081). A minor impact was observed on SSI rates and no impact was observed on CLABSI rates (Figure 1). This variation in percentage reduction for the different HAI types can be explained by how pathogens enter the host. CAUTI typically occurs as the result of pathogen contamination of either the internal or external surface of the urinary catheter^7^. Since the catheter can come in contact with a patient’s bed linen, any contamination on the bed linen can potentially be transferred to the external surface of the catheter, which provides a direct route of entry into the bladder where the pathogens can multiply and result in an infection.

**Figure 1.**
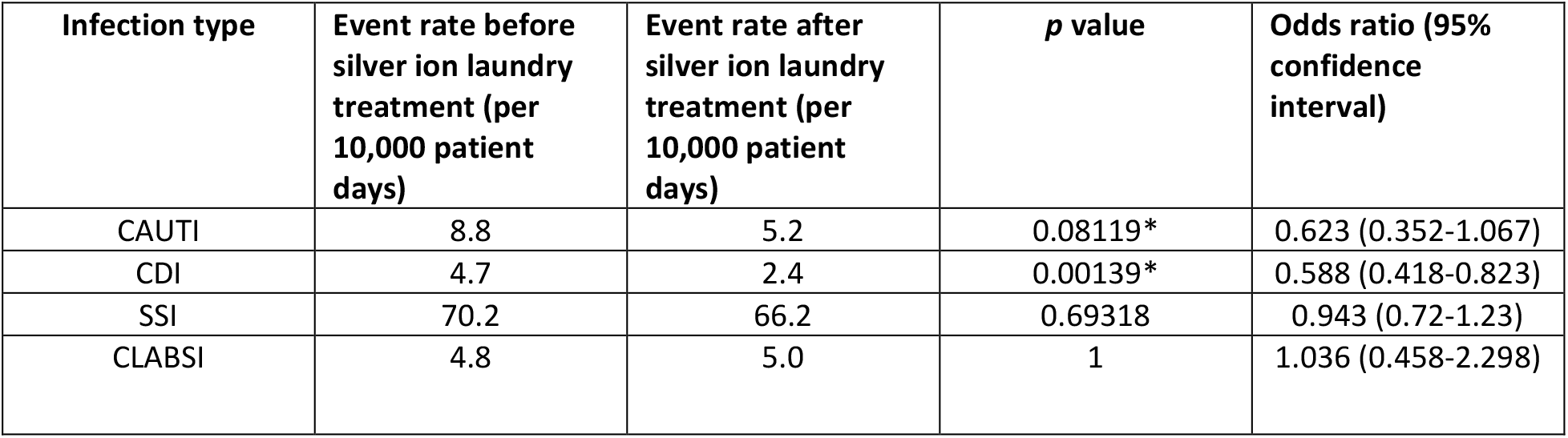
HAI rates (calculated per 10,000 patient days) at each of the three test hospitals demonstrated declines in the after-silver ion laundry treatment period. 30 months before silver ion laundry treatment period is 2013 Q3-2015 Q4; 30 months after silver ion laundry treatment is 2016 Q1-2018 Q2. * Indicates statistical significance

CDI typically is caused by the introduction of fecal bacteria to the patient’s mouth via contaminated hands or objects, usually at a time when the patient’s bowel flora has been disrupted by antibiotics and/or gastric ulcer prophylaxis^6^. Studies show that patients in rooms where an MDRO patient (such as CDI) resided previously will have an increased risk of infection due to insufficiently disinfected surfaces, including the floor^10^. Patients walking to and from the bathroom and back to bed could bring *C*.*difficile* from the floor, transferring it to the bed linen. From there it is possible for patient hands or objects to become contaminated, and then for that contamination to be transferred from hand to mouth, resulting in potential infection ^10^.

The impact on SSI was marginal. Possible explanations could include that infections are introduced during the procedure itself and/or that post surgically patients are on antibiotics. No impact was observed on CLABSI rates potentially indicating that textile-borne pathogens do not contribute to these types of infections. When the infection rates were evlauted for each individual hospital, consistent declines of 48%, 21% and 16% for each of the three hospitals (for a composite decline of 42%), primarily contributed by the declines observed in CAUTI and CDI rates (Figure 2). The hospitals observed a decline of 43% in overall infection rates at the interim 18-month evaluation period (8.72 to 4.91/10,000 patient days), during which time the control hospitals observed 4% decline in their infection rates (10.50 to 10.05/10,000 patient days). At this time, these hospitals dropped out as controls and began using the silver ion laundry treatment. While all three treatment hospitals showed a declining trend in infection rates, majority of the decline observed in the composite number was driven by Hospital 1, which also had the largest number of occupied beds.

**Figure 2.**
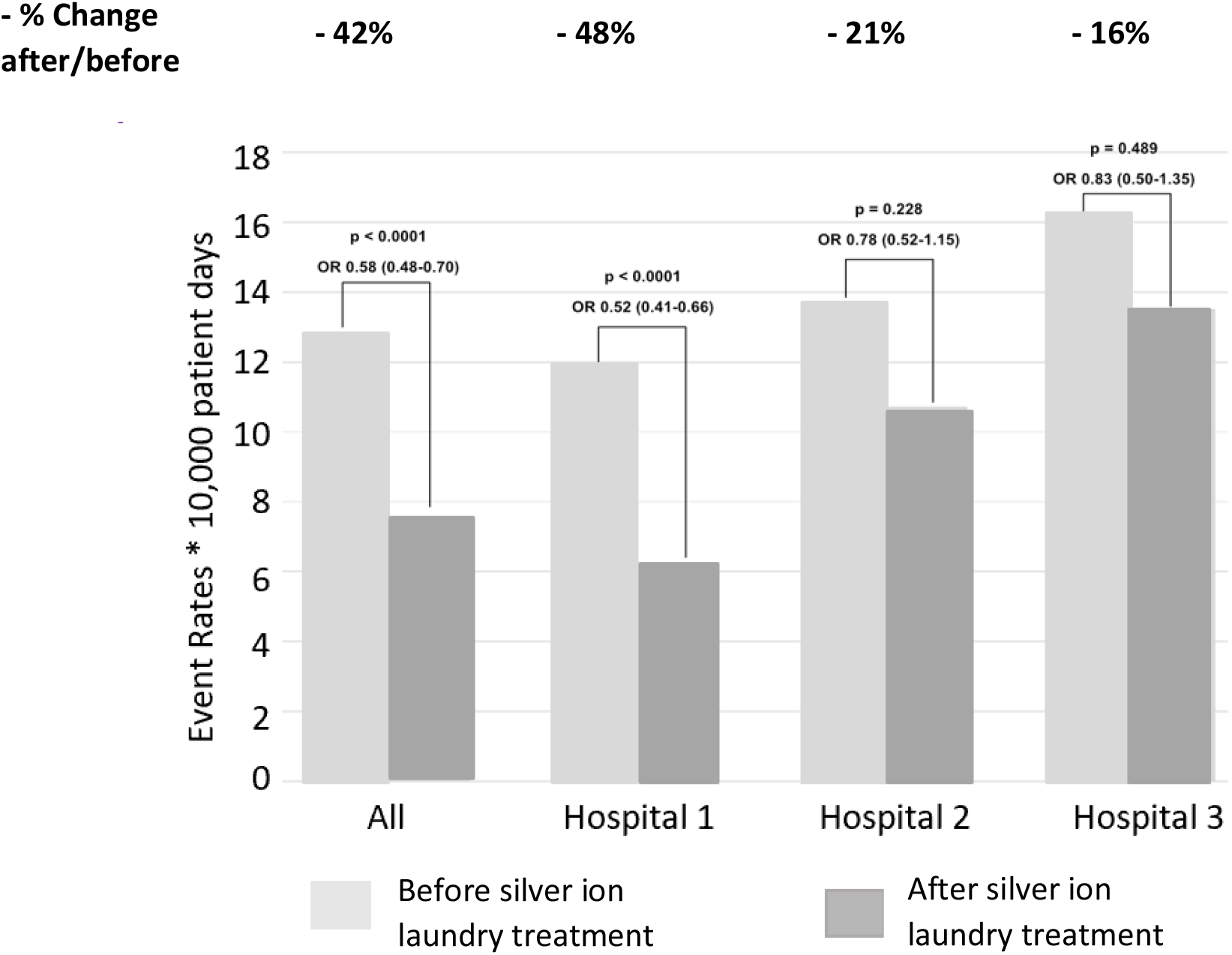
HAI rates (calculated per 10,000 patient days) at each of the three test hospitals demonstrated declines in the after-silver ion laundry treatment period. 30 months before-silver ion laundry treatment period is 2013 Q3-2015Q4; 30 months after silver ion laundry treatment is 2016Q1-2018Q2.

## CONCLUSIONS

This study highlights the benefit of a silver ion-based laundry treatment as an added element in infection prevention programs. This technology is fully automated, requires no additional inventory, training or change in workflow. The specific reduction in CDI and CAUTI reported in this study, suggests that certain infection prevention bundles may benefit more with this soft surface intervention than others.

One limitation of this study is that the exact contribution of the silver ion-based antimicrobial textile intervention cannot be measured using the retrospective anlayses study design. Additional studies would also be required to replicate these findings and to measure the impact of the laundry treatment on other HAIs (e.g. ventilator associated pneumonia), other high-touch surfaces such as privacy curtains, as well as complementarity to other infection prevention interventions. The combination of silver ion-based treatments’ ability to effectively combat both viruses and antibiotic-resistant pathogens makes it a potentially highly effective environmental infection control technology. Such technologies provide an important supplemental level of safety during infectious disease outbreaks and overall improvement in infection prevention programs.

## Data Availability

All data included in the manuscript is available.

## ACKNOWLEDGEMENTS

The authors acknowledge Elizabeth Hutt Pollard, Sean Morham, Sue Barnes, Bill Morris and Leo Selker for their considerable contributions.

